# Geographical distribution and the impact of socio-environmental indicators on incidence of Mpox in Ontario, Canada

**DOI:** 10.1101/2024.06.24.24309430

**Authors:** Chigozie Louisa J. Ugwu, Ali Asgary, Jianhong Wu, Jude Dzevela Kong, Nicola Luigi Bragazzi, James Orbinski, Woldegebriel Assefa Woldegerima

## Abstract

**Background:** Ontario, being one of Canada’s largest provinces, has been central to the high incidence of human Mpox. Research is scarce on how socio-environmental factors influence Mpox incidences. This study seeks to explore potential geographical correlations and the relationship between indicators of social marginalization and Mpox incidence rate in Ontario.

**Methodology:** We used surveillance data on confirmed human Mpox cases from May 1, 2022, to March 31, 2024, extracted from the Public Health Ontario website for this study. Spatial autocorrelation of Mpox incidence was investigated using spatial methods including Moran’s Index, Getis–Ord Gi*statistic, and spatial scan statistic. Following this, we adopted a generalized Poisson regression (GPR) model to estimate the incidence rate ratios (IRRs) based on the association between Ontario PHU-level marginalization and Mpox incidence, while adjusting for age and sex. The goodness-of-fit of the models was assessed using the Akaike Information Criterion (AIC), Akaike’s Information Criterion corrected (AICc), and the Bayesian Information Criterion (BIC).

**Results:** Spatial scan statistics, LISA, and Getis-Ord Gi*statistics revealed similar results for PHUs with the highest rates of Mpox in Ontario. Our study detected statistically significantly higher Mpox cases in Toronto, Ottawa, Peterborough, Kingston, Peel, Wellington-Dufferin-Guelph, Middlesex-London, Halton region, Brant County, Hamilton, and Haldimand-Norfolk PHUs (*p* − value < 0.05). Higher rates of Mpox infection in Ontario were associated with ethnic concentration (racialized, migrants or visible minority) (*RR* = 9.478; 95% *CI* = 1.621 − 2.876), and male gender (*RR* = 5.150; 95% *CI* = 1.159 − 2.119) and residential instability (*RR* = 14.112; 95% *CI* = 1.887, 3.407).

**Conclusion:** We identified major Mpox hotspots in Toronto. According to our model results, the high incidence rate may be influenced by the greater population of internal migrant population and younger individuals. Based on these insights, we recommend targeted interventions in the high-risk neighborhoods. Efforts to improve Mpox diagnosis and promote health equity among socioeconomically vulnerable populations, including racial and ethnic minorities, should be implemented.

## Introduction

In July 2022, the World Health Organization (WHO) declared the Mpox disease to be a pandemic due to its sporadic spread to non-endemic regions (1, 2). The global public health, social security, and the economy have been challenged by the Mpox outbreak in non-endemic areas, including Canada (1). In September 2023, the virus had infected over 90,000 people worldwide, and 117 countries had reported cases of deaths associated with the Mpox. Recent epidemiological studies from non-endemic countries have highlighted large differences in the incidence of Mpox in the context of geographical locations, population, and socio-environmental factors (3–6).

In Canada, the Mpox burden is unevenly distributed geographically across provinces and within provinces, while the highest number of cases were witnessed in Ontario (2, 7). As of September 31, 2023, there were a total of 1515 human Mpox cases in Canada and 770 human Mpox cases in Ontario alone, with no fatalities (1, 8).

Although vaccination and public health measures have been successful in reducing the spread of Mpox in Canada, the province of Ontario still observed an increase in Mpox occurrence from mid-January to the end of March 2024 with a total of 32 confirmed human Mpox cases, despite the decline across the country (9, 10). The new occurrence of Mpox cases suggest a possible disproportionate burden of confirmed human Mpox cases in Ontario in relation to socio-environmental and behavioral determinants (11).

In Canada, there is limited research on the role of social and demographic factors that may influence both the incidence and within province spread of Mpox (2, 9). This is an important area of study considering prior work conducted in Ontario in the context of similar diseases like Covid-19, where researchers found that people that are marginalized had increased risk of COVID-19 morbidity and mortality as compared to the general population in Ontario (12–14). This may demonstrate possible lack of health service equity (15).

It is epidemiologically recognized that socioeconomic status, marginalization, and inequity are significant risk factors for morbidity and mortality from various diseases (16, 17). Research has shown that a person’s socio-economic position, which includes factors like education, occupation, and income, can have a significant impact on their overall health (6). Typically, individuals with a lower socio-economic status are more susceptible to health issues and disease infection, hence, contributing to health inequities that impact individuals across all levels of society (1, 18).

In Ontario, the largest province in Canada, and most demographically diverse province in the country, vulnerability to disease infection could be influenced by the level of marginalization of the PHUs or neighborhood in which individuals live (19, 20). Although there is prior research on health equity and other diseases such as COVID-19 in the context of marginalization, there has not been any study on the role of social factors in the risk of contracting Mpox in Ontario.

Much work has been done in other non-endemic countries like the USA, European countries and in Brazil, with less literature in Canada (21, 22). Research evidence in other countries has shown that marginalized groups of individuals are more likely to be vulnerable to Mpox infection (6, 23, 24). Apart from sexually diverse groups, ethnic minorities and socioeconomically disadvantaged (material deprived) individuals may have disproportionately suffered socio-determinants of health, with disproportionate resource distribution, societal exclusion, and different life experiences having well-documented implications for health status and well-being (19). Understanding the epidemiological context, geographical distribution, and influence of socio-environmental indicators in Mpox incidence at PHU level in Ontario can be an effective strategy for planning public health policies and targeting priority areas.

Given the availability of the 2021 Ontario marginalized index data (25, 26), this study aims to examine the geographical distribution and the role of PHU-level marginalization indicators on the incidence of Mpox cases in Ontario, while adjusting for age and gender. We hypothesize that (1) Mpox infection rates are randomly distributed among the PHUs in Ontario (values of the features i.e. Mpox incidence rates throughout Ontario are spatially uncorrelated) and (2) PHUs with higher population density and greater levels of marginalization are likely to experience an increased incidence of Mpox. This study could guide resource allocation, including testing, vaccination strategies, and other disease control interventions, to Public Health Units (PHUs) at higher risk of Mpox.

## Materials and methods

### Study area and data

The study region, as shown in **Fig 1**, is the province of Ontario in Canada, which is partitioned into 34 units called Ontario Public Health Units (PHUs). Ontario is a highly populated province in the east-central part of Canada. Ontario has an estimated population of 14.2 million, a land area of 892,411.8 *km*^2^with a population density of 16 inhabitants/*km*^2^ (27). Approximately 34% of the population are visible minorities, and about 44% are immigrants born outside Canada (28).

**Figure 1.**
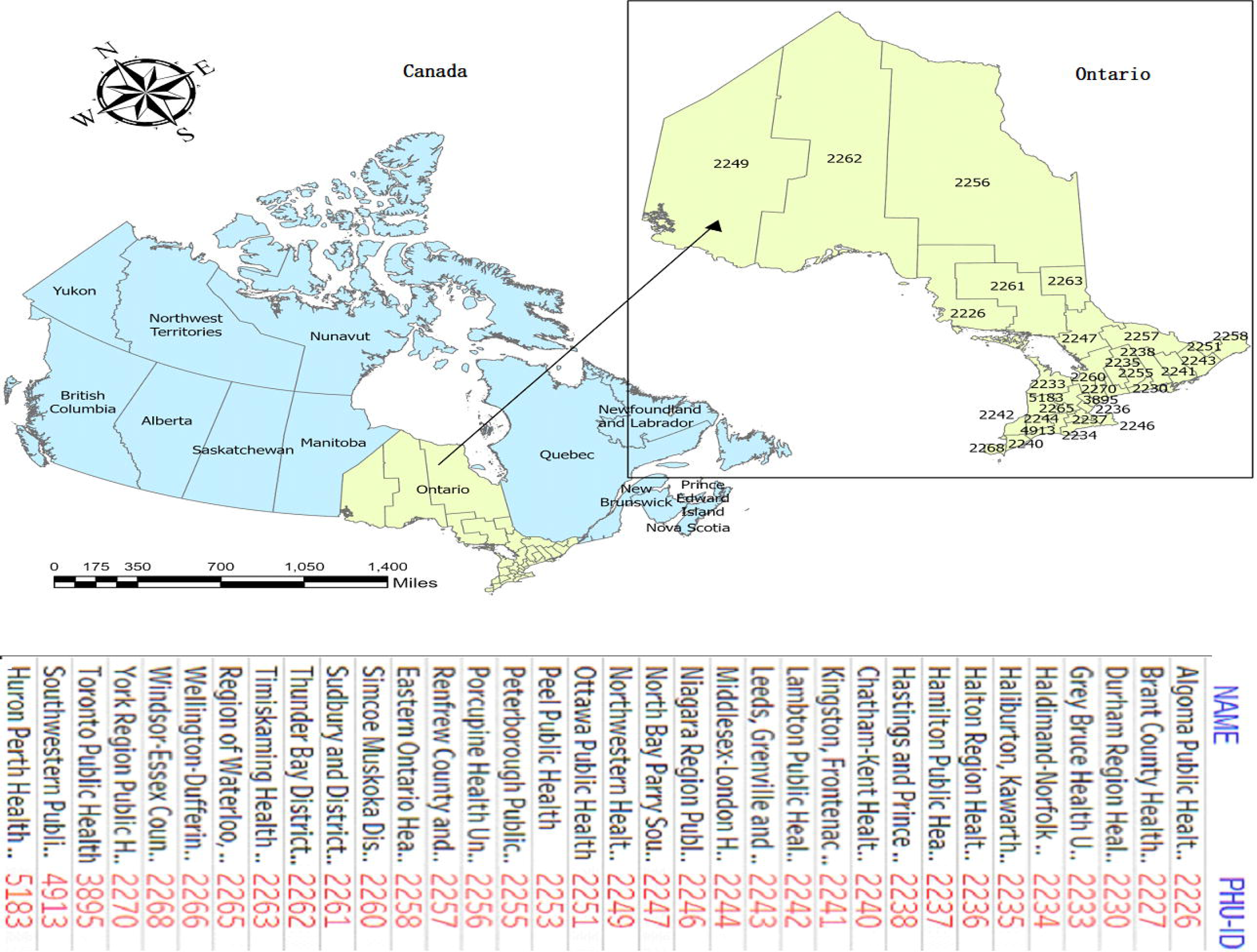
Geographic location of the study area. Ontario is in east-central Canada and the country’s second largest province by land area in km^2^.

The PHUs, which were formed by grouping several urban and rural municipalities, are the smallest geographical units considered for this study (28). Algoma (2226), Northwestern (2249), Thunder Bay District (2262), Sudbury and District (2261), and the Timiskaming PHUs are situated in Northern Ontario, while the remaining 29 PHUs make up Central, Toronto, Southern and Eastern Ontario Health Unit Regions (28).

### Mpox epidemiological data

We obtained data on confirmed human Mpox cases in Ontario from May 1, 2022, to March 31, 2024, from the Integrated Public Health Information System (iPHIS) of Public Health Ontario (PHO), which records cases of reportable Mpox disease across Ontario and publish as surveillance report (9, 10). Between May 1, 2022, and July 2023, Ontario reported a total of 722 confirmed Mpox cases, with no deaths associated with the disease (11). Notably, there was a resurgence in Mpox cases beginning in mid-January 2024, with 32 confirmed cases reported by the end of March 2024, slightly fewer than the 33 cases recorded throughout 2023 (9). Overall, our study included a total of 758 Mpox cases across Ontario’s PHUs. We utilized the 34 PHUs (*n* = 34), as the spatial unit of analysis. All Mpox cases were geographically linked to the corresponding PHU using boundary polygons in ArcGIS Pro (29). These PHU-level polygons (geographical shapefiles) were sourced from the publicly accessible Statistics Canada Geodatabase (28, 30).

### Population and socio-demographic data

Based on existing studies and data availability, our explanatory variables included a variety of socioeconomic, and demographic factors initially retrieved at PHU level from the 2021 Statistics Canada population census data (30, 31). Apart from the population density, age distribution and gender, these variables included: (1) percentage of the population aged 15–64 years with a lower level of education (not having a university certificate, diploma or a bachelor degree), (2) prevalence of low income (living in a low-income household based on based on the Low-income measure, after tax (LIM-AT), table representing the poverty line), (3) unemployment rate (population over 15 years and unemployed), (4) percentage of immigrants (individuals who were born outside of Canada), and (5) average household size.

However, we found that the listed variables from 1 to 5 are subset variables used to construct the 2021 Ontario Marginalized index (ON-Marg) factors, a widely used index that encompass various factors of socioeconomic and marginalization status at Ontario PHU level (25, 26). The 2021 ON-Marg Index was created through principal component analysis from the 2021 Canadian census data, jointly by researchers at MAP Centre for Urban Health Solutions at St. Michael’s Hospital and Public Health Ontario (25).

These are four dimensions (subdomains) of marginalization as measured by the 2021 ON-Marg for each PHU in Ontario and have been used extensively across Ontario for research purposes on health and disease disparities, advocacy work, population health assessment and surveillance, and public health program planning and resource allocation (25, 32). Therefore, as described in **Table 1**, this study extracted the 2021 ON-Marg index (26), for each PHU in Ontario, derived using principal component factor analysis of 42 indicators from the 2021 Census of Population (33, 34). Additionally, the four-dimensional ON-Marg factors have been demonstrated to be linked to many health outcomes and preferred for use in this research to overcome the issue of multicollinearity, and to demonstrate its potential to measure health inequalities in the context of Mpox incidences in Ontario.

**Table 1:** Explanatory variables.

| Variable | Description |
| --- | --- |
| **Material resources or deprivation** | The ON-Marg dimension of material deprivation is closely **connected to poverty (low socio-economic status)** and analytically factored from the 2021 population census data using these indicators:<br>(1) Proportion of the population aged 15 years + without a high-school diploma (low level of education).<br>(2) Proportion of families who are lone parent families.<br>(3) Proportion of the population aged 15 years+ who are unemployed.<br>(4) proportion of the population considered low-income households (prevalence of low-income), and<br>(5) Proportion of households living in dwellings that need major repair. |
| **Population density** | Percentage of people residing in each PHU per km/square (measurement of population per unit land area for each PHU). |
| **Household dwelling (residential instability)** | Composite measure of:<br>(1) Proportion of population living alone.<br>(2) Average number of individuals per household (dwelling)<br>(3) Proportion of the population aged 15+<br>(4) Proportion of population who are single, divorced, or widowed. |
| **Racialized and newcomer populations (ethnic concentration or visible minority)** | Composite measure of:<br>(1) Proportion of population who are recent immigrants (within 5 years), and<br>(2) Proportion of the population who self-identified as a visible minority (as defined by Statistics Canada), they are and/or non-white, non-Indigenous populations. |
| **Age and labour force (dependency)** | Composite measure of:<br>(1) Proportion of the population who are 65+<br>(2) Proportion of population not participating in labour force (aged 15+)<br>(3) Dependency ratio (total population aged 0 to 14 and 65+ over the total population aged 15 to 65 years. |
| **Gender** | The proportion of males aged 15+ of the total population of males and females residing in each PHU. |
| **Percentage age 0-14 year** | Age groups of the population at each PHU from 0-14 years |
| **Percentage age 15-64 year** | Age groups of the population at each PHU from 15-64 years |
| **Percentage age 65 year and over** | Age groups of the population at each PHU from 64 years and over. |
| **Average household size** | The average number of persons per household. |
| **Married or common law partner** | Proportion of married or common law partners who set up their household together in one dwelling. |
| **For each dimension, scores were treated as a continuous variable from lower to higher level of marginalization** |  |

These dimensional factors (**Table 1)** included: material resources (previously called ‘material deprivation’), households and dwellings (previously called ‘residential instability’), age and labour force (previously called ‘dependency’) and racialized and newcomer populations (previously called ‘ethnic concentration’) or Percentage of visible minorities, have been associated with many health outcomes (25, 26).

According to the Employment Equity Act, visible minorities are “persons, other than Aboriginal peoples, who are non-Caucasian in race or non-white in colour”. We also adjusted for demographic factors such as gender, and population density, and age distributions by PHU extracted from the 2021 Canadian census (31). Population density was measured as the number of persons per square kilometer, and it was found to be highly skewed to the right and therefore, a log-transformation was applied.

Datasets were linked to the geographic boundary files, sourced from Statistics Canada and Ontario Open Data (31), using the PHU-ID in Arc GIS Pro. (29)

### Ethical considerations

The current study utilized publicly available Mpox data; therefore, ethical approval is not required. The data is available online at: https://www.publichealthontario.ca/

### Measurement of a dependent variable

The Mpox incident rate per 100,000 population for the years 2022-2023 was used as the dependent variable. To calculate this rate, we employed the 2021 census population data to ascertain the population at risk of Mpox for each PHU in Ontario, using the formula:

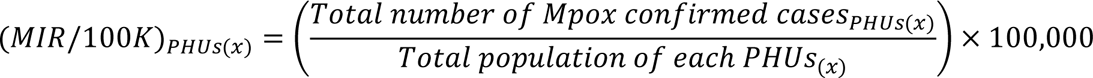

Subsequently, the Mpox incidence rate per 100 000 population for each PHU and the explanatory variables were linked to the PHU polygon in the ArcGIS Pro version 3.0.36056 for further analysis (35).

## Analytical methods

### Global spatial autocorrelation analysis of Mpox incidence rate

This study first applied spatial autocorrelation to determine whether Mpox cases are correlated geographically. Spatial autocorrelation, as determined by Global Moran’s Index (36), acts as the indicator for analyzing Mpox geographical distribution throughout Ontario. Spatial autocorrelation is closely related to Tobler’s first law of geography, which states that “everything is connected to everything else, but objects at proximity are more strongly interconnected than those farther apart.” (37). A Global Moran’s Index statistics equal to zero (0) shows the absence of spatial correlation, indicating a random distribution of Mpox, and no clustered PHUs across Ontario. A Global Moran’s Index > 0, indicates the presence of positive spatial autocorrelation, and when the value approaches +1, it signifies a strong positive spatial autocorrelation, indicating that the PHUs are clustered.

In this study, we evaluated the spatial autocorrelation of the Mpox incidence rate throughout Ontario using the global Moran’s I, and the interpretation of the result was considered in the context of its null hypothesis of spatial randomness (36). The null hypothesis specifies that the Mpox incidence are randomly distributed among the PHUs within Ontario. A *p* − *value* resulting from the Global Moran’s I test below the 5% significance level indicates the presence of spatial autocorrelation of Mpox among the PHUs across Ontario. Let, *x*_*i*_ and *x*_*j*_ denote observed Mpox cases at PHUs *i* and *j*, *i*, *j* = 1,…, *n* = 34. Then the global Moran’s I statistic (36), is defined as:

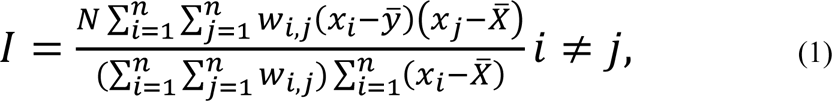

where *N* is the total number of Mpox cases, *n* is number of PHUs, *W*_*i*,*j*_ denote the spatial weights between PHU *i* and PHU *j*; 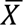 represents the mean value of Mpox cases across the entire PHU and it is given by 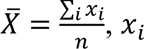 and *x*_*j*_ denote the number of Mpox cases in PHU *i* and *j*, respectively,. The corresponding values of *z* −score and *p* − *value* of Moran’s I statistic are used to reject or accept the null hypothesis of spatial randomness of Mpox distribution across the entire PHUs in Ontario. For this study, we employed a fixed distance band spatial matrix for the autocorrelation (Global Moran’s I) analysis.

### Local indicators of spatial autocorrelation

#### Local Moran’s I statistic

The major limitation of Global Moran’s I statistic is that it only generates a single summary statistic for the entire study area, meaning that it only measures spatial autocorrelation (clustering) at the global scale (38). Global Moran’s I does not detect where local clusters or spatial outliers are located (39). Therefore, we applied the local Moran’s I statistic (LISA) introduced by (39), to investigate the local level of spatial clustering of PHUs with a high and low incidence of Mpox (38). The computation of local Moran’s I assesses the local version of Global Moran’s I for each PHU, computes scores that reveal the underlying significant spatial clustering and local spatial outliers within the data at each PHU to determine variation in spatial autocorrelation over Ontario. Its significance is evaluated in five categories namely: High-High, High-Low, Low-High, Low-Low, and non-significant Mpox incidence rates (40). LISA effectively pinpoints PHUs where Mpox incidences are significantly notable, offering insights into potential underlying mechanisms [39]. It calculates a local Moran’s I value, a z-score, a pseudo p-value, and a code representing the cluster type for each statistically significant PHU. The z-scores and pseudo p-values indicate the statistical significance of the computed Local Moran’s I values (39), at *p* − *value* < 0.01.

Let *x*_*i*_ be the *ith* Mpox observation at the *ith* PHU, then, the local Moran’s I statistic (41) of spatial association is calculated using the following formula:

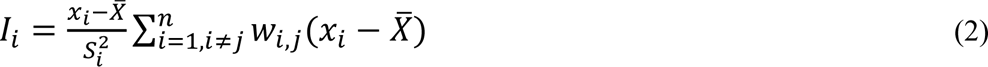

where *x*_*i*_ is the Mpox incidence rate in the *ith* PHU; *w*_*i*,*j*_ is the spatial weight matrix that defines spatial interaction between PHU *i* and *j*; *n* is the total number of PHUs, 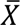 is the mean and 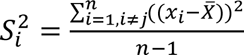 is the deviation of neighbouring PHUs (39). This study employed the K-nearest neighbors’ approach to establish a spatial weight matrix, wherein the spatial autocorrelation relationship was defined with 33 neighboring Public Health Units (PHUs) for each specific PHU (41). We used ArcGIS Pro (ESRI, Redlands, CA, USA) (35), LISA analysis. The statistical significance level was set at 0.05, and we used 9999 permutations in the simulation to assess the sensitivity of our results (41).

#### Getis-Ord local Gi* statistic

As alternative to local Moran’s I inferential statistic, local Getis-Ord Gi* statistic was applied to pinpoint more local level statistically significant Mpox hotspots, cold spots and relative magnitude in each PHU (42). While a PHU with a high Mpox count is noteworthy, it may not necessarily be a statistically significant hotspot. Unlike the local Moran’s I, the Getis-Ord Gi* statistic value computed in each PHU is expressed as z-score value, which allows a direct interpretation for statistical significant of the PHU (42). The Getis-Ord Gi* statistic is computed using the following formula:

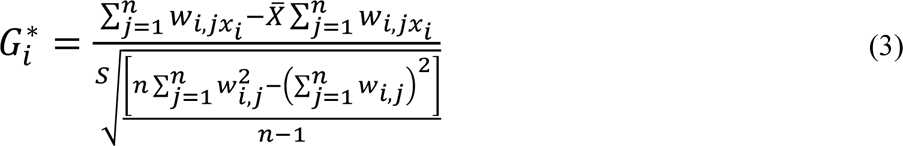

where *x*_*i*_ is the Mpox count at *ith* PHU, *w*_*i*,*j*_ is the spatial weight between *ith* and *jth* PHU, *n* denote the number of PHUs, 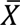 is the mean and *s* is the standard deviation. We used ArcGIS Pro (ESRI, Redlands, CA, USA) (35), the Getis-Ord Gi* statistic, setting the number of permutation tests at 9999 for sensitivity analysis and a 0.05 level of significance. The K-nearest neighbors’ approach was utilized, with contiguity set at 34 PHUs for a polygon contiguity spatial weight matrix. This matrix was created based on the 34 PHUs that share common boundaries and vertices. (41).

#### Spatial scan statistics

We utilized a spatial scan statistic (43), with a discrete Poisson probability model to evaluate the spatial distribution of Mpox incidence in Ontario, as LISA and Getis-Ord Gi* statistics only consider neighboring Public Health Units (PHUs). This analysis aimed to identify statistically significant spatial clusters of Mpox, both high and low, for comparison with Mpox hotspots identified by the Getis-Ord Gi* statistic (44). Unlike the LISA and Getis-Ord Gi* statistics, the retrospective purely spatial scan Poisson model detects most likely high clusters accurately because it fits the assumption that the number of Mpox cases in each PHU was Poisson distributed according to a known underlying population at the risk of Mpox.

The analysis employed a circular moving window centered on each PHU, with a maximum spatial cluster size based on the population at risk of Mpox. The window moved until reaching the maximum population at risk, with the radius of each circle continuously increasing to a maximum radius. The maximum likelihood function was used to maximize the circular window size, identifying the primary cluster as the circular window with the highest likelihood ratio (44). Additional non-overlapping clusters in circular windows with high likelihood ratios were identified as secondary clusters. Each circular window was tested using a Monte Carlo simulation to assess the null hypothesis of spatial randomness. SaTScan version 10.1.3 was used for this analysis, with a maximum window size of 50% of the population at risk (45). To ensure statistical power, 999 Monte Carlo replications were performed, considering only clusters at a 99% confidence interval as statistically significant.

### Non-spatial statistical analysis

Descriptive statistics and bivariate association between Mpox outcomes and explanatory variables were explored. To test for global spatial dependency, we calculated the Global Moran’s I, and no statistically significant spatial autocorrelation was detected with the Global Moran’s Index. Thus, we proceeded with the multivariate Poisson analysis without accounting for spatial correlation (46). The final model choice was guided by the Akaike Information Criterion (AIC), Akaike’s Information Criterion corrected (AICc), and the Bayesian Information Criterion (BIC) (47).

We considered using the Poisson Regression (PR) model as the initial statistical model in the count families of generalized linear models (GLM) to model our data (48). Let *y*_*i*_ be the Mpox count variable in the *ith* PHU, *i* = 1,2,…,34. If *y*_*i*_∼ a Poisson distribution, the probability density function is expressed mathematically as:

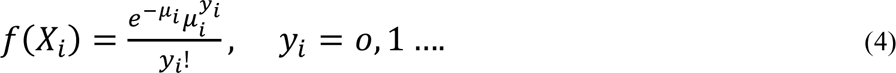

The Poisson model assumes that the mean equals the variance, *E*(*X*_*i*_) = *var* (*X*_*i*_) = μ_*i*_ =*exp exp* (*X*_*i*_). To integrate a covariate into Equation (4) while ensuring non-negativity, the mean is presumed to be multiplicative, such that: *E*(*X*_*i*_) = μ_*i*_ = *X*_*i*_ *exp exp* (*X*_*i*_β), where *φ*_*i*_ is a measure of disease exposure, *X*_*i*_ is a *p* × 1 vector of predictor variables (socioenvironmental variables), and β is a *p* × 1 vector of unknown regression coefficients.

The regression coefficients (β) are estimated via the maximum likelihood approach by maximizing the log-likelihood function. The Poisson log-likelihood is given as:

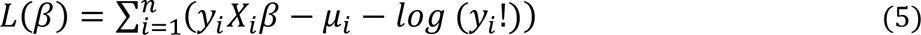

Even though the standard PR model has advantages, it also has potential drawbacks. The application of the standard PR model often encounters violations of the assumptions of independence and excessive variance (i.e. Over dispersion, fitted variance being larger than the mean), which requires model adjustment (49). The problem of adjusting for overdispersion in the standard PR model is that it leads to inflated test statistics, biased standard errors, and inconsistent conclusions of estimates. The presence of over-dispersion can be assessed by comparing the deviance 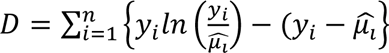, with the degree of freedom (48).

Corresponding to the outcomes being over dispersed count data, a generalized Poisson regression model (GPR) was chosen as a flexible extension to the standard Poisson regression model (50). The GPR model proposed by (51), accommodates both over-dispersion, under-dispersion and cluster heterogeneity inherent in a data (52). The GPR has a probability density function is given as (51):

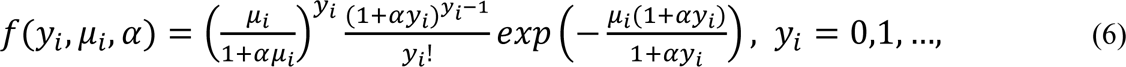

If α = 0, the GPR model reduces to standard Poisson regression, resulting to *E*(*y*_*i*_) = *var*(*y*_*i*_). The mean of the GPR model is given as *E*(*y*_*i*_|*x*_*i*_) = μ_*i*_ and variance *var*(*y*_*i*_|*x*_*i*_) = μ_*i*_/(1 + αμ_*i*_)^2^. Clearly, when α > 0, the variance is over-dispersed, and when 2/μ_*i*_ < α < 0, the variance is under-dispersed (51). The maximum likelihood estimates of the GPR parameters 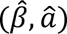 can be computed by maximizing the log-likelihood of GPR

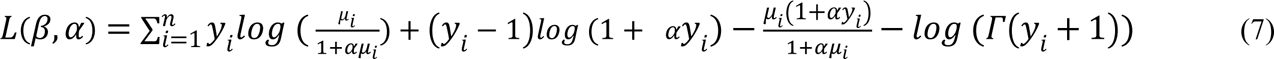

The Newton-Raphson numerical iteration approach is mainly used to maximize the log-likelihood function, where the first and second derivatives of the log-likelihood are required (51). The sequential iteration procedure is implemented to obtain the maximum likelihood estimates 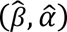.

#### Model comparison and fitting

In this study, two possible count models were considered, namely, the standard Poisson Regression and its generalized Poisson Regression (GPR) model. To distinguish the most suitable and well-fitted count regression model for the Mpox data, Akaike Information Criterion (AIC), Akaike Information Criterion corrected (AICc), and Bayesian Information Criterion (BIC) were used (53). After the model comparison, the GPR model was fitted to our data to model the impact of the marginalized index variables including, material deprivation, ethnic concentration, dependency ratio and residential instability on Mpox cases at the PHU level with population as an offset, while adjusting for age and gender (51). All statistical analyses were conducted using SAS (version 9.4, Cary, NC, USA) (54), and displayed as incidence rates of IRRs for Mpox with 95% CIs. Computed 95% CIs that do not include unity were considered statistically significant.

## Results

### Descriptive results

A total of 758 Mpox cases were notified in Ontario from 2022 to March 2024. **Fig 2** and **Fig 3** display the distribution of Mpox cases by age group and gender, respectively. The highest Mpox infection rate is among individuals aged 30–39 years, accounting for ∼ 67% of Mpox cases (**Fig 2**). This age group had the highest or similarly high rates of Mpox infection across all public health units. The data also indicates that males are more frequently affected by Mpox infections than females (**Fig 3**). As evidenced in **Fig 4**, the bar chart distribution of Mpox incidence rates across Ontario indicated that the highest Mpox incidence rates were recorded in Toronto as compared to other PHUs.

**Figure 2.**
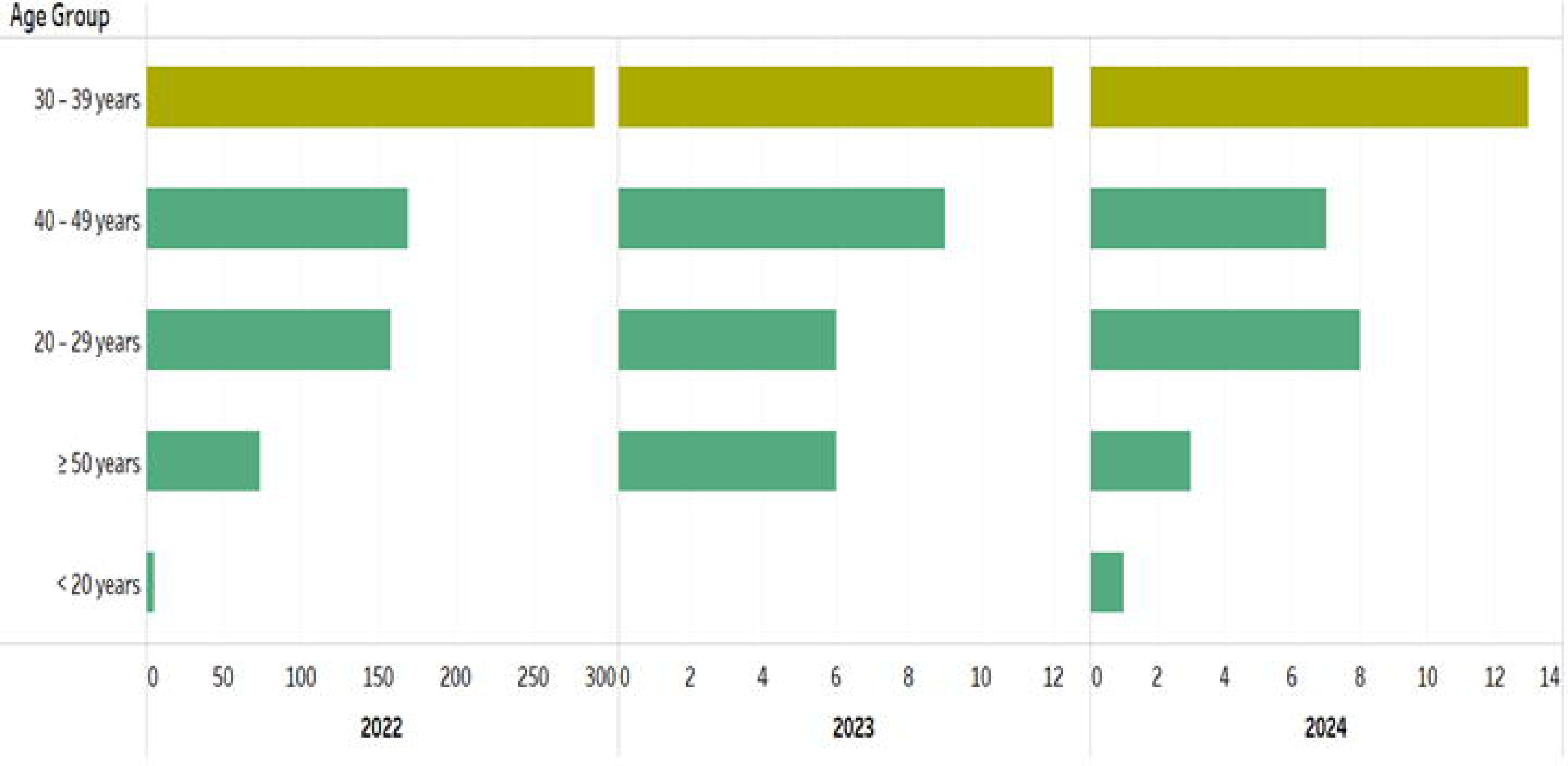
Distribution of Mpox by age group (**2022–2024**).

**Figure 3.**
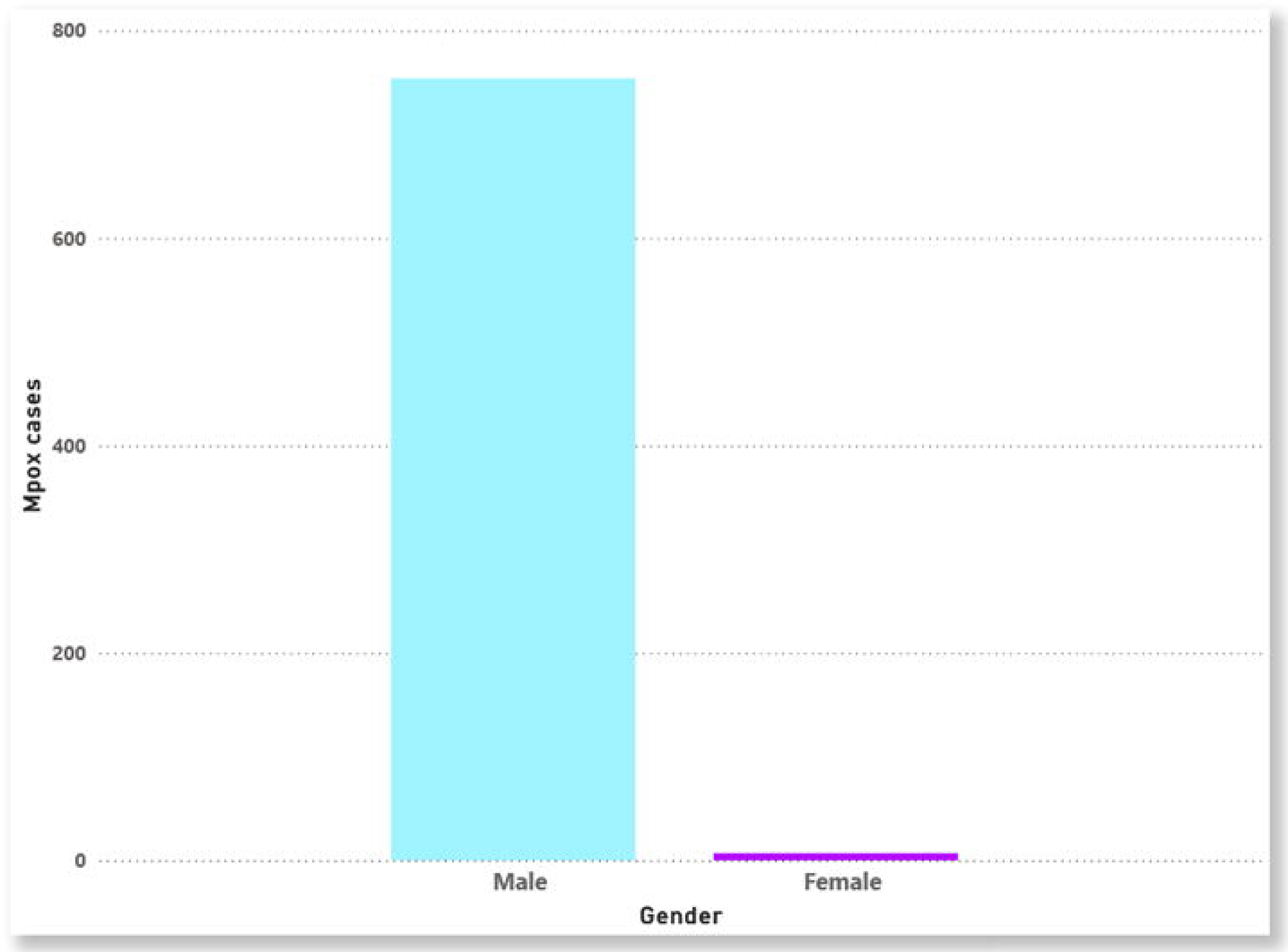
Distribution of Mpox by gender (**2022–2024**).

**Figure 4.**
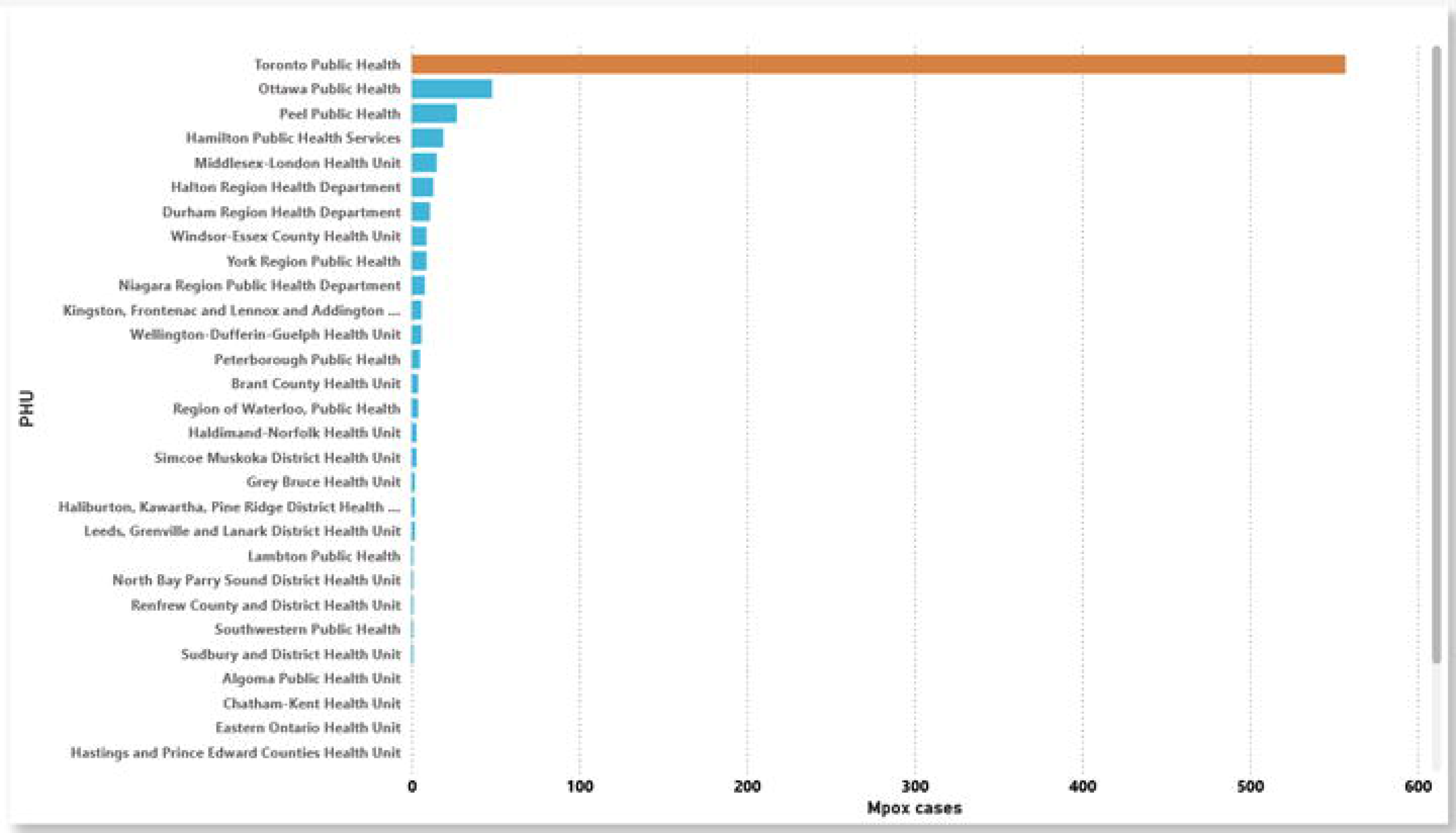
Bar chart of Mpox cases across the Ontario PHUs.

### Spatial analysis results

The Choropleth map in **Fig 5** displays the geographical distribution of the Mpox incidence rate in Ontario at the PHU level. Seven of the 34 PHUs have incidence rates for Mpox greater than 2.6 per 100, 000 population, including Toronto (3895), Ottawa (2251), Peterborough (2255), Hamilton (2237), Kingston (2241), Brant County, and Middlesex-London (2244) PHUs, all of those from Eastern, Central-east, Central-west and South-west health unit regions. Another seven of the remaining 27 PHUs have incidence rates for Mpox greater than 1.2 per 100, 000 population including Wellington-Dufferin-Guelph (2266), Peel (2253), Halton region (2236), Windsor-Essex County (2268) Haldimand-Norfolk (2234), Durham (2230), and Niagara region (2246) PHUs, five of those from Central-west health unit region. Other PHUs had considerably low and no Mpox incidence rate was recorded in the North-west and North-east health unit regions (**Fig 5**).

**Figure 5.**
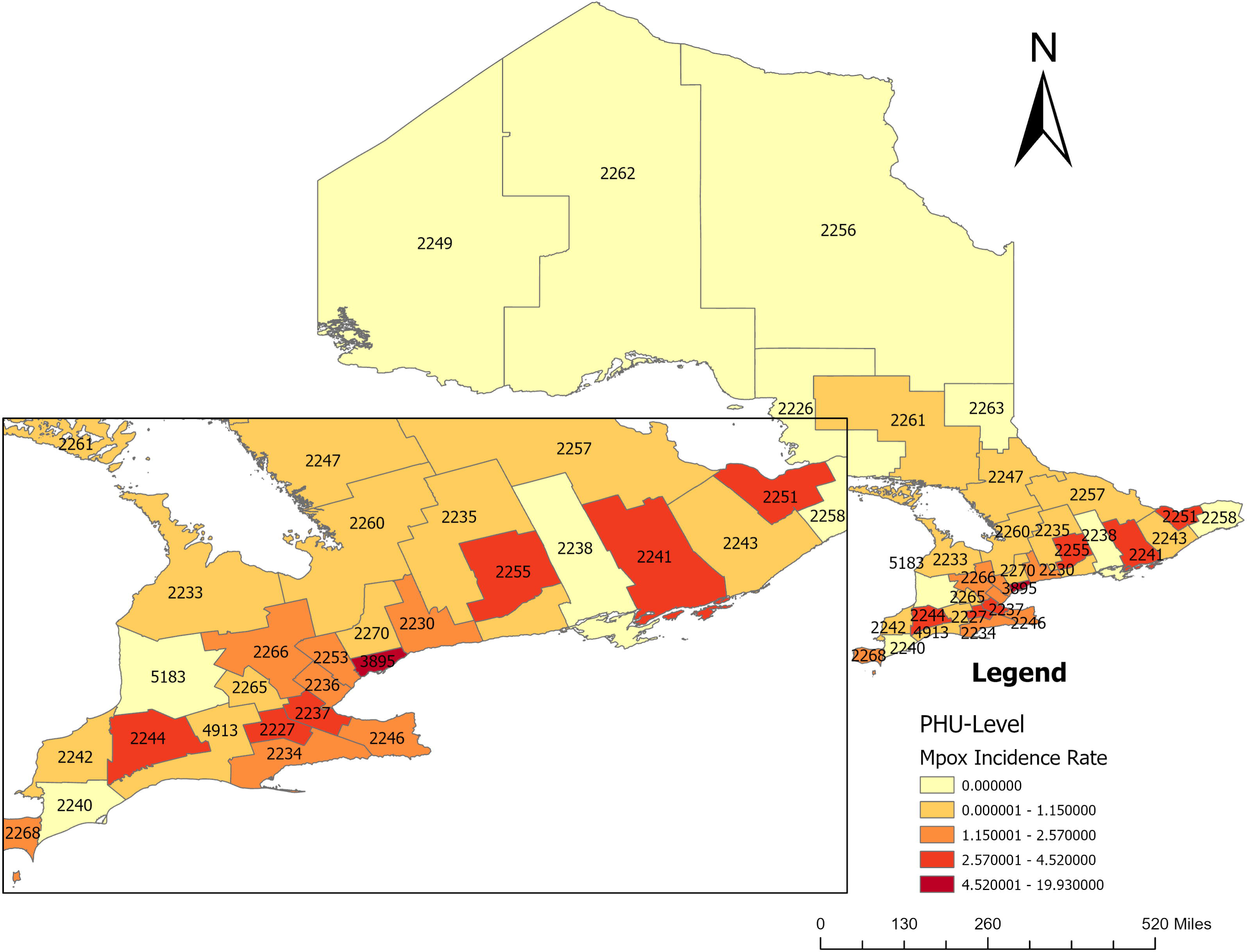
Choropleth map of geographical distribution of Mpox incidence rate in Ontario (Incidence rate: case per 100,000 population) (PHU names are available in Fig 1).

The global Moran’s Index value was (*Moran’s I* = 0.025079, *z* − *score* = 1.433116, *p* − *value* = 0.151825 > 0.05) for Mpox incidence rate as shown in **Fig 6**. The result of the global Moran’s I indicated no presence of statistically significant spatial autocorrelation in Mpox incidence rate over the whole study region (Ontario PHUs). Given the z-score of 1.433116, the Mpox spatial pattern does not appear to be significantly different than random, meaning that the Mpox incidence rate in the model is randomly distributed at the PHU level. The specific value is displayed in **Fig 6**. Therefore, applying spatial smoothing in this context would be misleading, as it assumes a spatial correlation does not exist (41).

**Figure 6.**
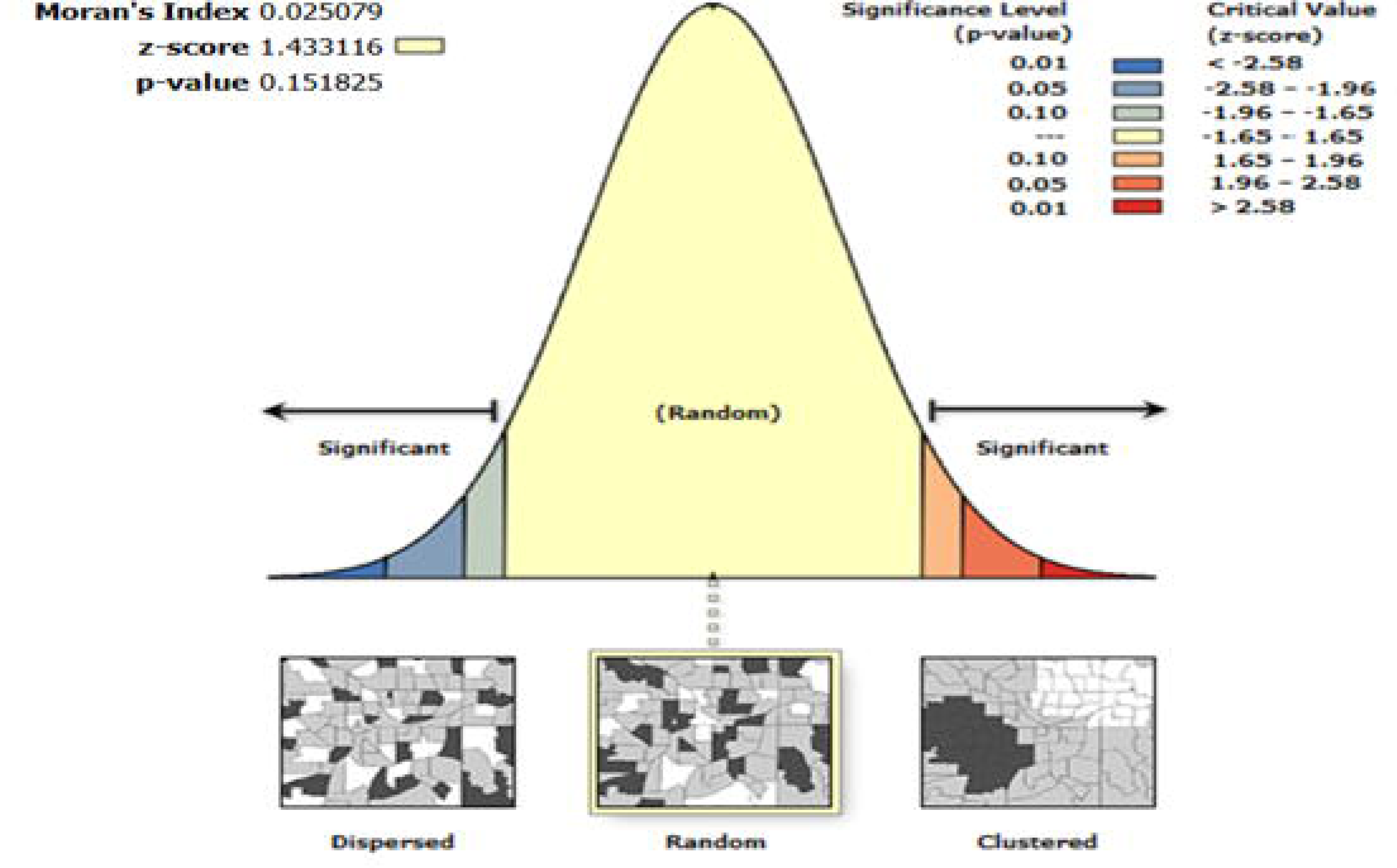
Global spatial autocorrelation analysis of Mpox incidence rate in Ontario (global Moran’s I).

The Local Moran’s I statistic indicated that several Public Health Units (PHUs) in Ontario, including Toronto (3895), Peterborough (2255), Kingston (2241), Ottawa (2251), Peel (2253), Wellington-Dufferin-Guelph (2266), Middlesex-London (2244), Windsor-Essex County (2268), Halton Region (2236), Brant County (2227), Hamilton (2237), and Haldimand-Norfolk (2234), exhibited statistically significant High-Low Outliers for Mpox incidence throughout the study period **(Fig 7).**

**Figure 7.**
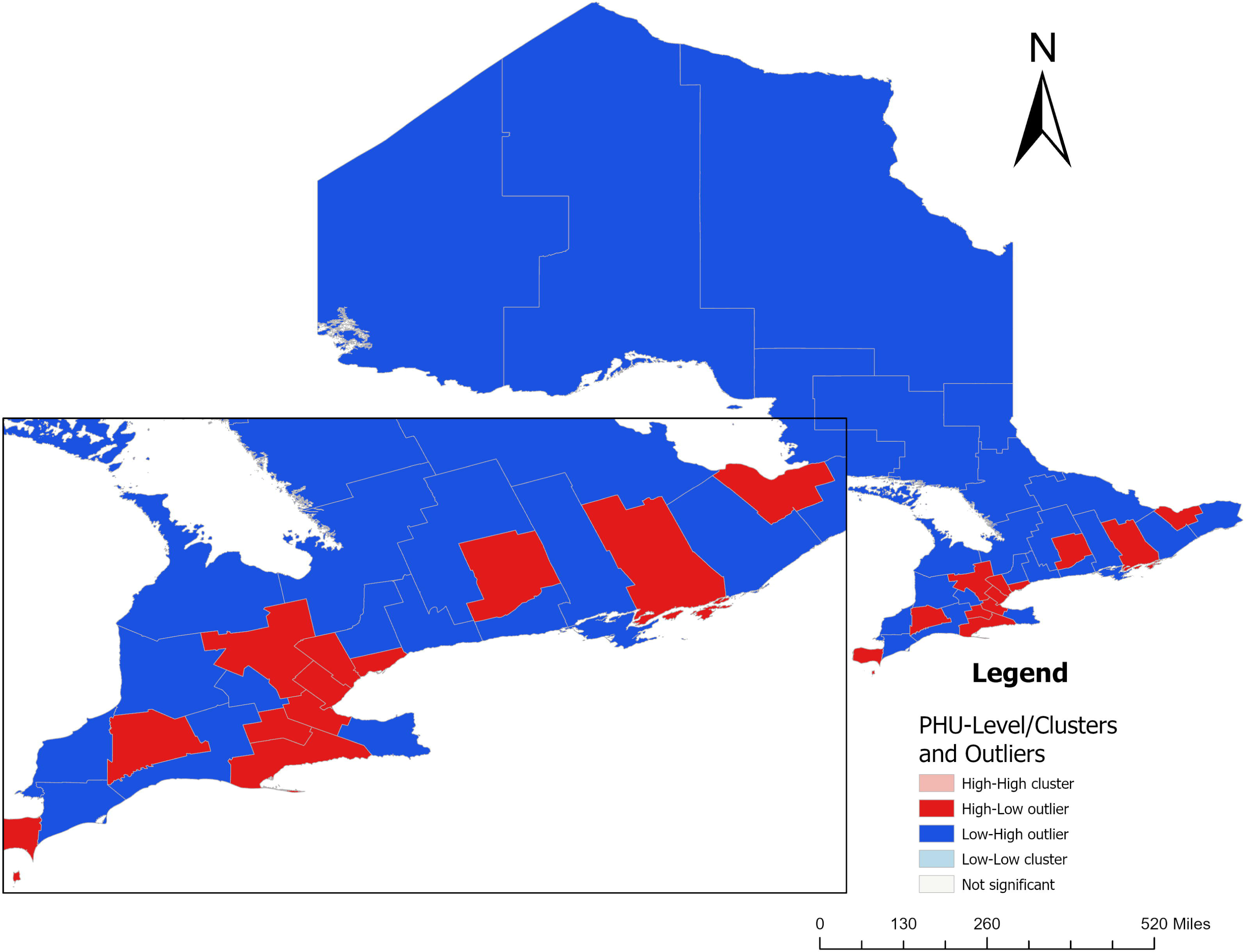
Local spatial autocorrelation analysis of Mpox incidence rate in Ontario (local Moran’s I) (PHU names are available in Fig 1).

**Fig 8** presents a hotspot analysis for Mpox in Ontario using the local Getis-Ord Gi* statistic. The map clearly shows that Mpox incidence hotspots are primarily located in Toronto, with a 99% confidence level of significance. However, the cluster overlaps with neighboring PHUs, namely Durham Region (2230), Peel (2253), and York Region (2270), which have a statistically significant level of 90% (**Fig 8**). These results suggest that Toronto has neighborhoods with a high incidence of Mpox, potentially due to sociodemographic and marginalized index factors. Therefore, targeted outreach programs focusing on Mpox screening interventions and resource allocation are recommended.

**Figure 8.**
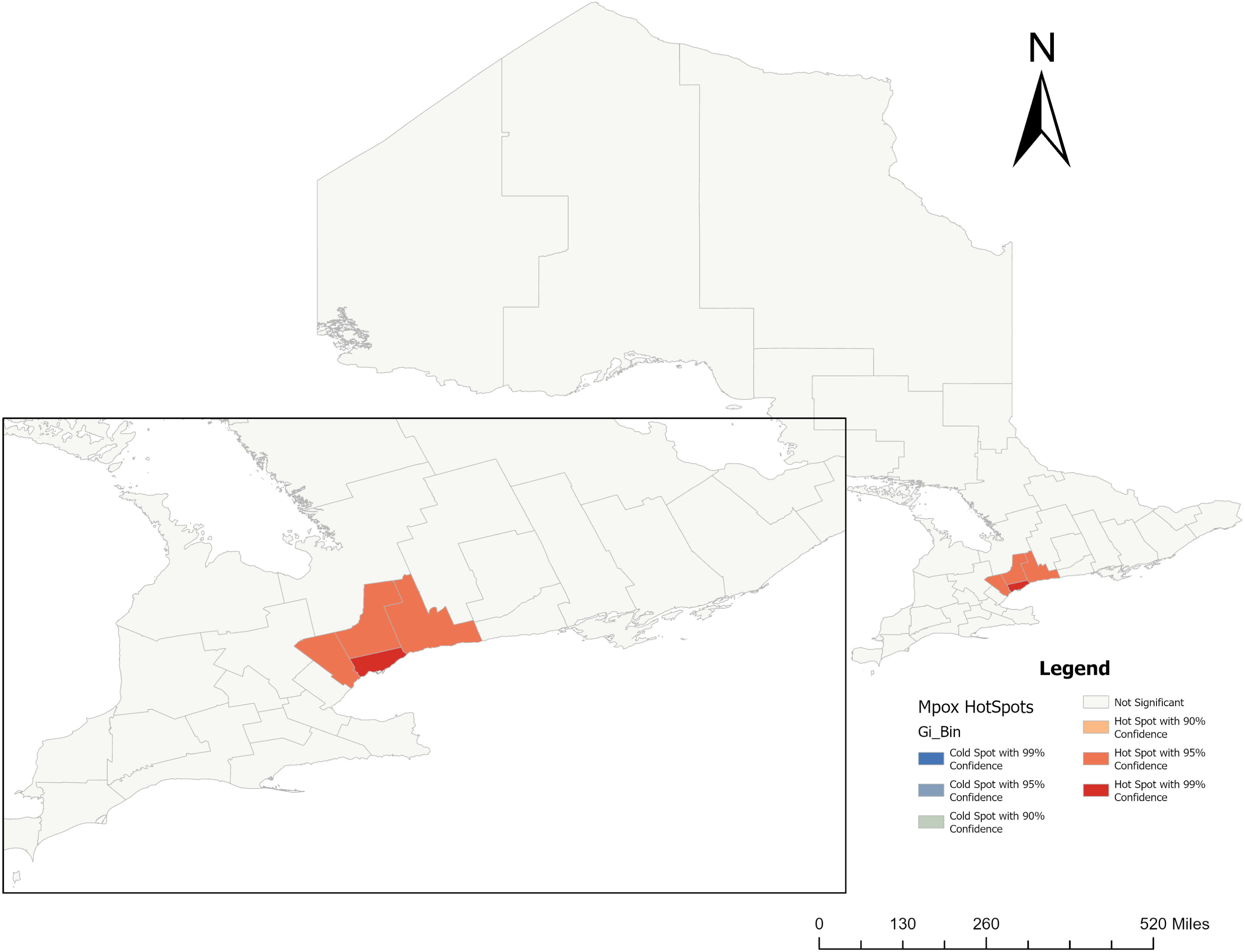
Getis-Ord Gi* hotspot analysis of Mpox incidence rate in Ontario (PHU names are available in Fig 1).

The spatial scan statistic revealed two spatial clusters (**Fig 10 and 11**). The primary cluster was located in one PHU (**Toronto**). There were 557 Mpox cases compared to 149 expected cases. Thus, the ratio between the observed and the expected Mpox cases was 3.74. The p-value was 0.0001, smaller than 0.05 significant level, which indicated that the cluster was highly significant. The relative risk (RR) for the population inside the cluster compared to the population outside the cluster was 11.34, indicating that the risk of Mpox within Toronto was higher than locations outside it (neighboring PHUs). The RR is estimated risk within the cluster divided by the estimated risk outside the cluster. The Log likelihood ratio of the primary cluster is 511.975.

**Figure 9:**
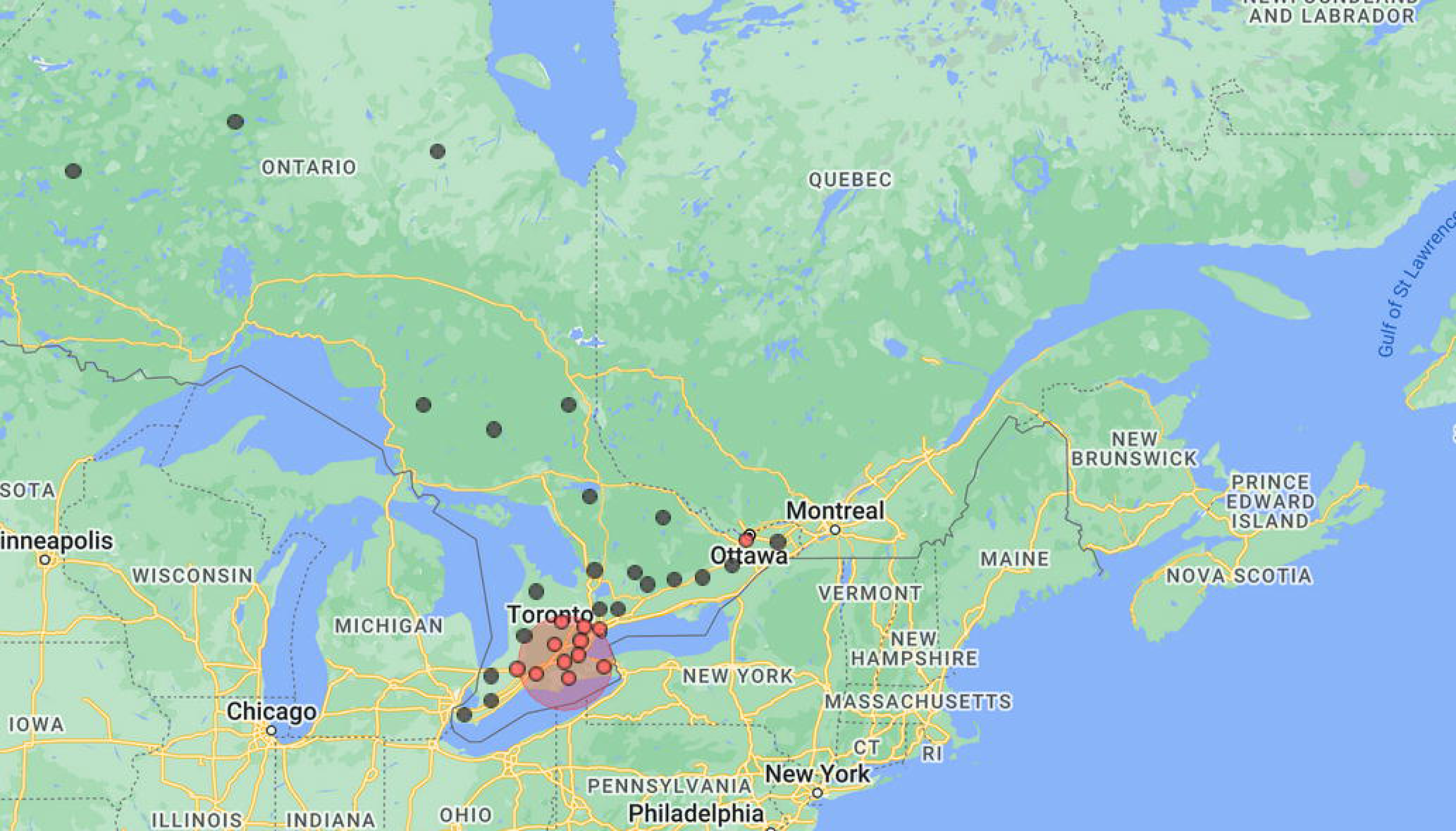
Spatial scan statistic calculated in SaTScan to detect significant Mpox local clusters.

**Figure 10:**
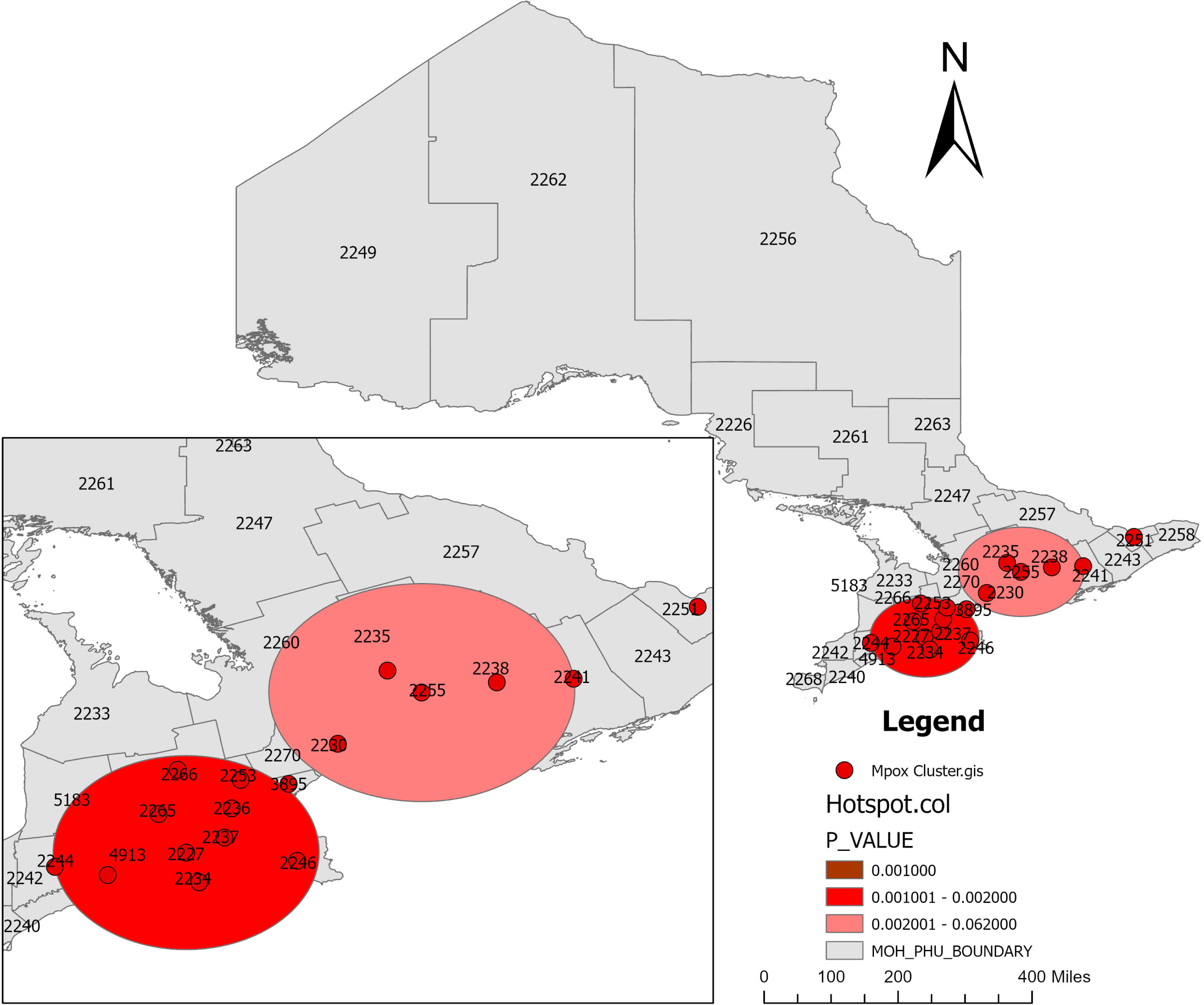
Spatial scan statistic calculated in SaTScan to detect significant Mpox local clusters (visualized in ArcGIS Pro).

**Figure 11:**
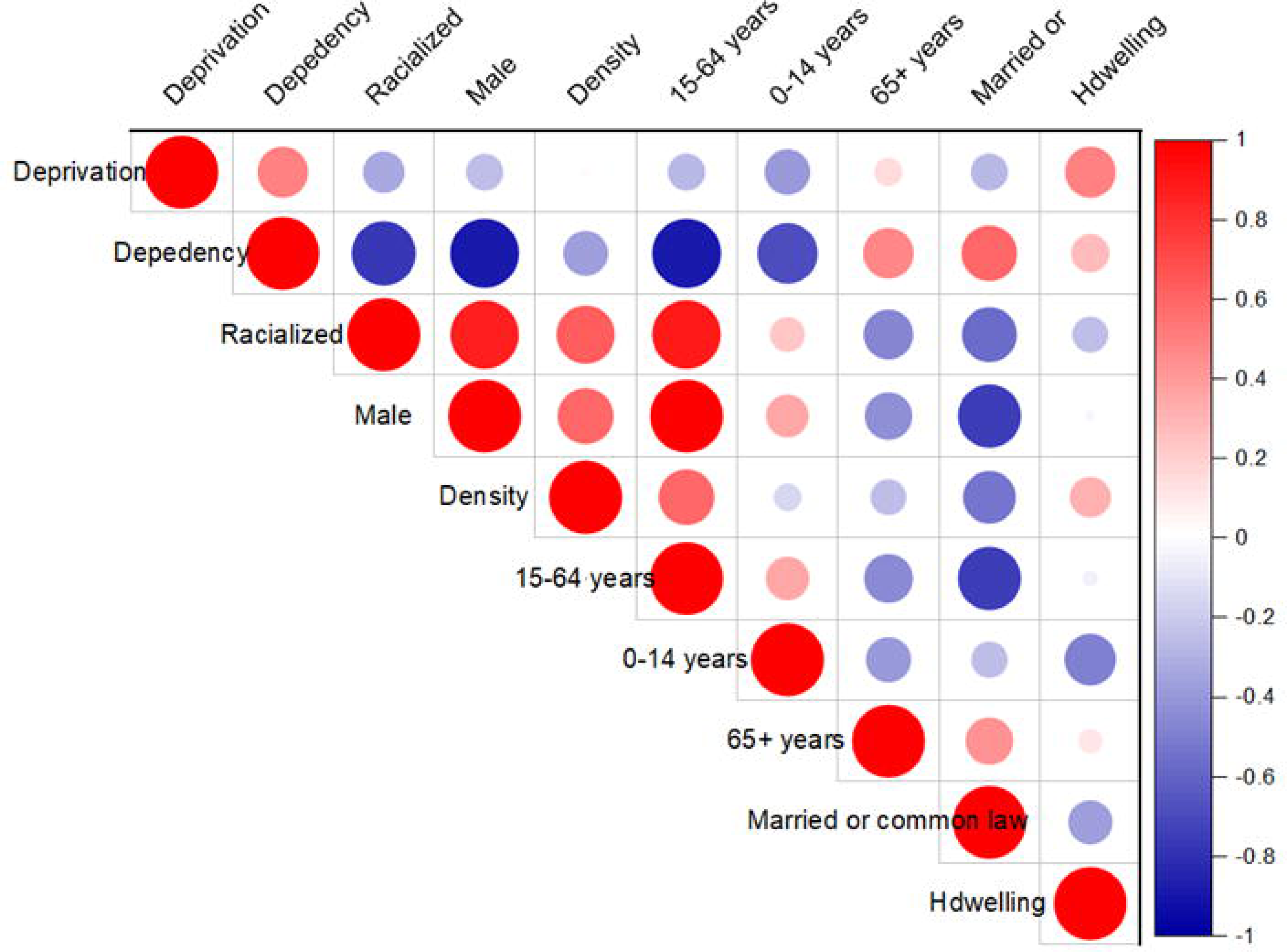
A Pearson correlation matrix illustrates the interdependence of the explanatory variables. Correlation coefficient values may be negative (−1) or positive (+1). If the correlation value is less than zero, it is weak, if it is larger than zero.

The spatial scan analysis also revealed that two secondary cluster, which is the most likely clusters, were located (a) in one PHU (Ottawa), and (b) overlapped by Toronto and Hamilton PHUS (the overlapped cluster is made up of districts in Kitchener, Brampton, Guelph, Mississauga, Brantford, and London). The first secondary cluster (a) has 48 number of observed Mpox cases was, compared to 17.89 expected Mpox cases. Thus, the ratio between observed and expected cases was 2.68. According to these calculations, the relative risk (RR) inside the cluster was 3.21, while the Log likelihood ratio was 19.884. The p-value was smaller than 0.05, indicating that the cluster was highly significant. The relative risk for the population inside the cluster compared to the population outside the cluster was 3.21, indicating that the risk of Mpox in Ottawa PHU was higher than locations outside the PHU (Fig **9 and 10**).

For the second secondary cluster (b), which is made up of nine local districts, the number of observed Mpox cases was 100, compared to 73 expected Mpox cases. Thus, the ratio between the observed and expected Mpox cases was 1.37. According to these computations, the RR inside the cluster was 2.07, while the Log likelihood ratio was 9.649. The p-value was 0.0004, smaller than 0.05 level of significance, indicating that the cluster was also significant. The relative risk for the population inside the cluster compared to the population outside the cluster was 2.-7, indicating that the risk of Mpox in these areas was higher than locations outside the areas (**Fig 9 and 10**).

### Non-spatial statistical analysis results

Initial Global Moran’s I test for spatial autocorrelation (**Fig 6**) indicated an absence of spatial correlation, suggesting that spatial dependence does not need to be considered in the data. However, before fitting the data, we examined the multicollinearity among the variables described in **Table 1**, as well as the confounding risk factors using a Pearson’s correlation analysis (**Fig 11**), because the presence of multicollinearity among explanatory variables can cause overfitting and less reliable inferences about the associations between the response and predictor variables in (55). We removed variables that were found to be highly correlated with ON-Marg dimensional factors and included other cofounding risk factors not correlated. Simultaneously, the variance inflation factor (VIF) was used to verify multicollinearity. The VIF values for all the filtered variables were less than 5, indicating no serious multicollinearity exists (55). This means that all the predictor variables in the final model were not highly correlated to each other.

Through the test of overdispersion in the standard Poisson regression analysis, we detected a potential problem of overdispersion. The deviance value was 30.017 and the scaled Pearson Chi-Square (*X*^2^), by degrees of freedom was 54.169, with *p* − *value* < 0.001, much bigger than one. In **Fig 12**, it was shown that the dependent variable (Mpox) is over-dispersed and has excessive zeros.

**Figure 12.**
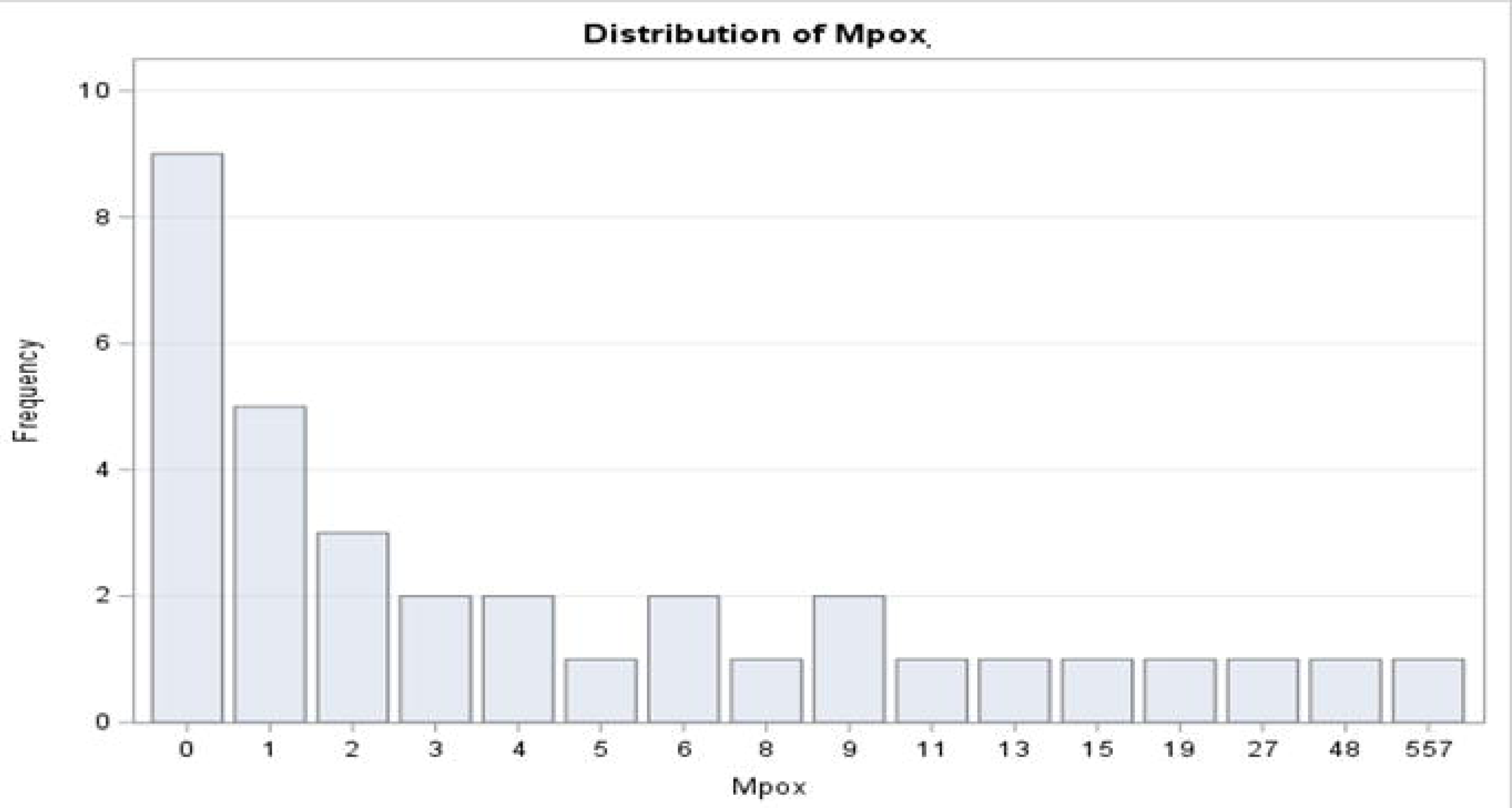
Histogram of distribution of Mpox counts in Ontario PHUs.

These findings indicate that the standard PR model does not fit our data well. PR modelling of data with overdispersion can lead to underestimation of standard errors, resulting in biased estimates of regression coefficients and misleading conclusions (48). One possible approach is to apply the generalized Poisson regression (GPR) model (51). In **Table 2**, AIC, AICc and BIC results also showed that the GPR model offers a better fit compared to the standard Poisson regression model and study was interpreted based on the results obtained from the GPR model.

**Table 2.** Model comparison (Criteria for Assessing Goodness of Fit).

| Description | AIC<br>(smaller is better) | AICC<br>(smaller is better) | BIC<br>(smaller is better) |
| --- | --- | --- | --- |
| Standard Poisson | 178.8521 | 189.1598 | 201.3582 |
| GPR | 163.6763 | 183.5366 | 174.3608 |

The standard Poisson regression results in **Table 3** demonstrated significant p-values for all variables, however, based on the result of model comparison (**Table 2**), we fitted the GPR model to handle the issue of overdispersion inherent in our data and to estimate accurate/unbiased incidence rate ratios (IRRs) with the corresponding 95% confidence intervals (CIs).

**Table 3:**
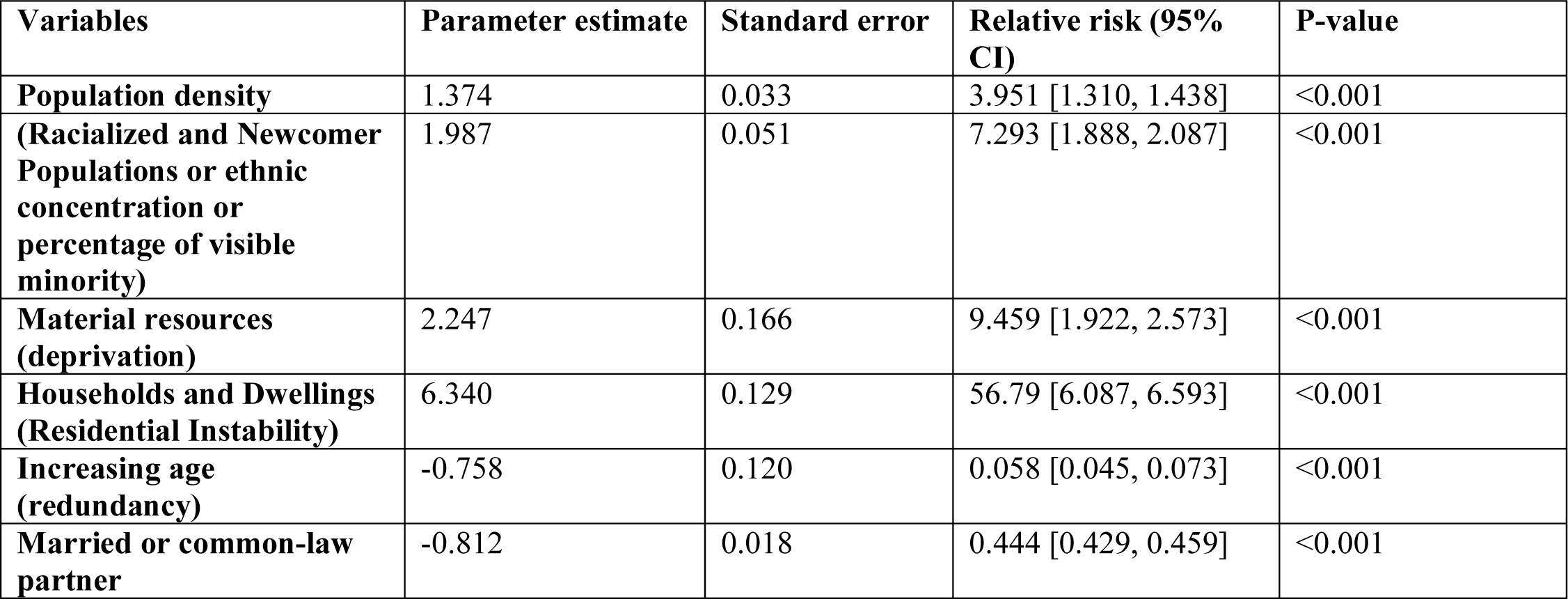
Standard Poisson regression coefficients for factors affecting Mpox counts in Ontario, at PHU level.

The GPR model was fit (**Table 4**), however, the population density was found to be correlated to proportion of racial population (ethnic concentration), and married or common-law partners in the correlation structure of the GPR and we used only the proportion of racial population to represent both population density and married or common-law partners. After removing two of the correlated variables, the final GPR model found only two of the ON-Marg factors the proportion of racialized population (newcomer /ethnic concentration or a visible minority) (*RR* = 9.478; 95% *CI* = 1.621 − 2.876), residential instability (*RR* = 14.112; 95% *CI* = 1.887 − 3.407), and male gender (*RR* = 5.150; 95% *CI* = 1.159 − 2.119) to be significant predictors of Mpox in Ontario.

**Table 4:**
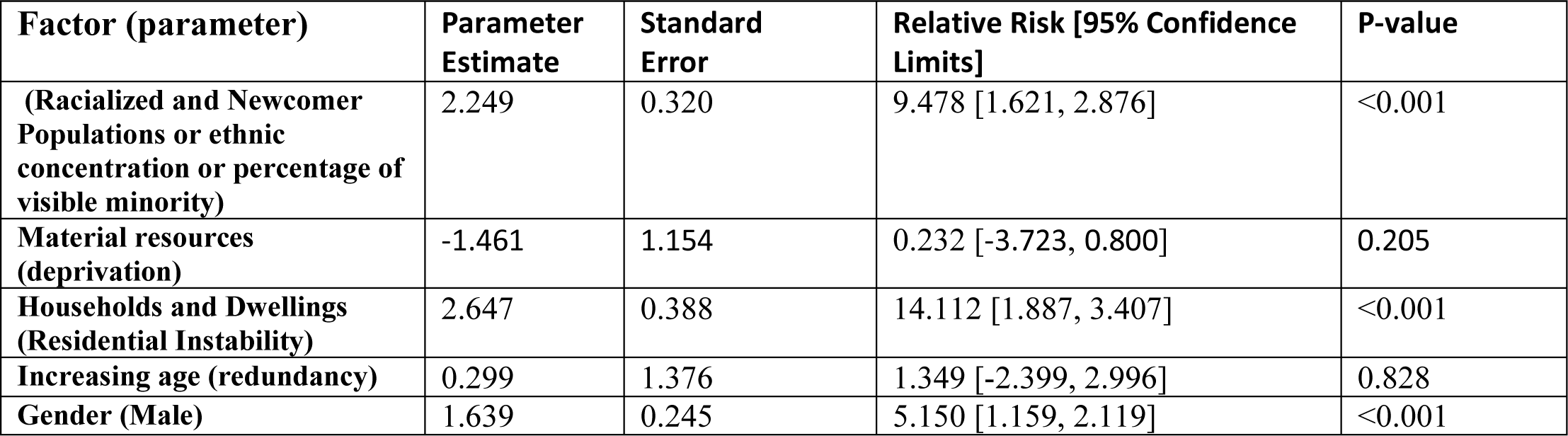
Generalized Poisson Regression coefficients for factors affecting Mpox counts in Ontario, at PHU level.

#### Significant interaction effects

PHU-level specific associations between Mpox occurrence and socioenvironmental factors were examined modeling an additional interaction term. The interaction effects (**Fig 13**) show a clear increasing trend in the rate of Mpox infection as population increased. It is worth noting that those living in the most ethnically diverse (racial population) have a considerably higher rate of Mpox (*p* − *value* < 0.001). The result indicated that increasing population density and proportion of these visible minority groups statistically significantly increased the risk of Mpox in high-risk areas (*p* − *value* < 0.001). Interestingly, as age increases the risk of Mpox in the population decreases (*p* − *value* < 0.001), and as material deprivation increases in a lower population density, Mpox infection also increases (*p* − *value* < 0.001).

**Figure 13:**
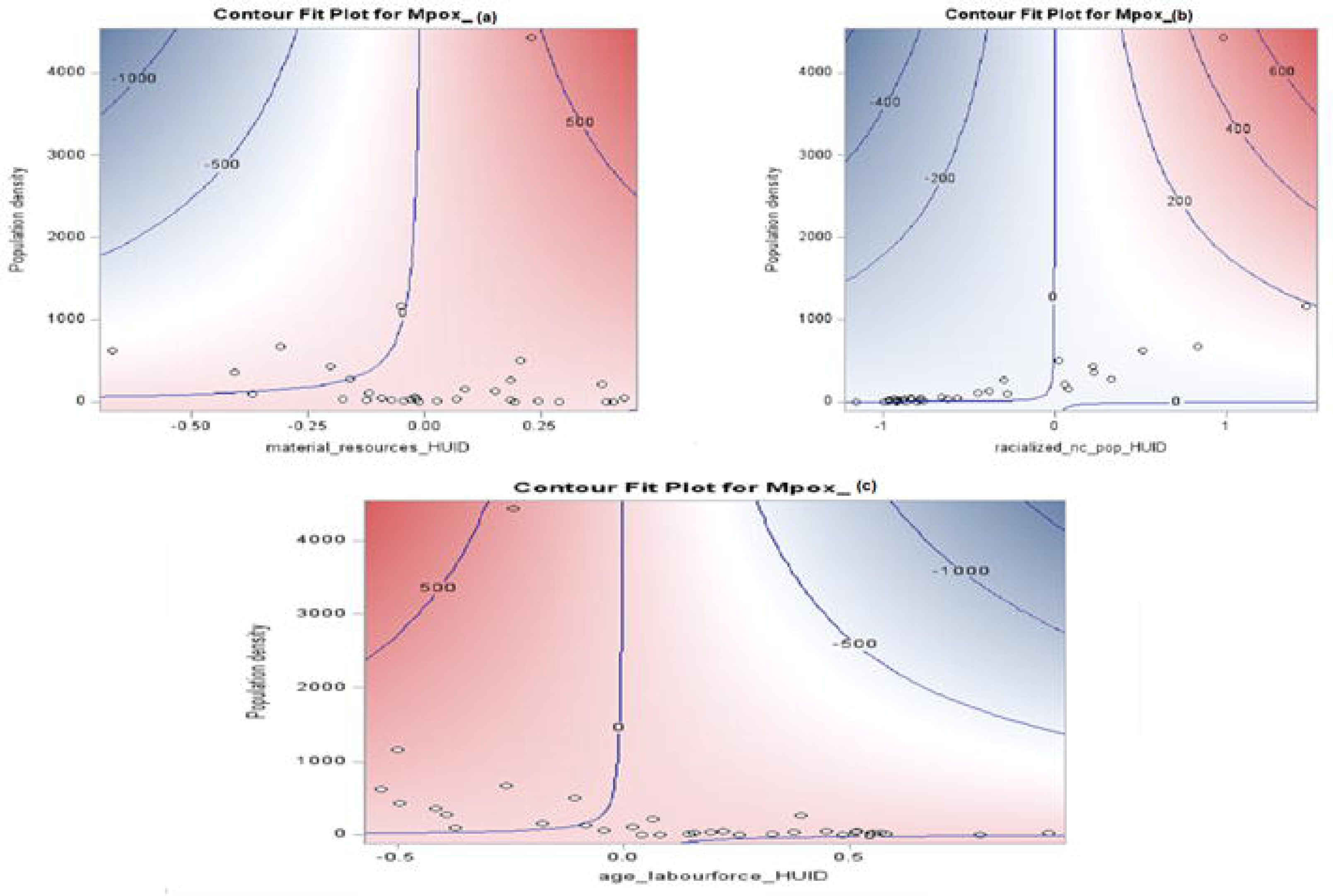
Interaction effect of population density, material deprivation and Mpox (a); population density, ethnic concentration and Mpox (b); population density, age and Mpox (c). The Mpox outcome is plotted as the contours. The blue areas denote lower counts of Mpox, while the redder areas denote higher counts, and the contours are labeled with the corresponding Mpox counts.

## Discussion

In this study, we observed that Mpox incidence rates exhibited a random spatial distribution pattern across the Public Health Units (PHUs) in Ontario based on Global Moran’s Index statistic. However, we found that the risk of contracting Mpox was higher in specific PHUs and was associated with population density as well as part of area-level marginalization. The spatial scan statistics, LISA, and local Getis-Ord Gi* statistics revealed similar results for areas with the high rates of Mpox. However, the clusters identified by the spatial scan statistics covered a wider area of Ontario districts and detected secondary and overlapping clusters. In contrast, the clusters identified by LISA and the local Getis-Ord Gi* statistic were more dispersed. Notably, the primary Mpox hotspot area was detected in Toronto, this is an area marked by substantial internal presence of migrant population (ethnic concentration or visible minority). Specifically, Toronto has a population of ∼2.8 million, with socioeconomic factors (25). The use of spatial correlation methods was effective in this study, as it enabled the identification of priority PHUs to address Mpox vaccination strategy, intervention, and resource allocation deficiencies in the province of Ontario.

Our study used the uniquely available 2021 marginalization index variables to evaluate the association between marginalization and overall Mpox incidence across the PHUs in Ontario using generalized Poisson regression. Based on literature, there may be a possibility of marginalized areas in both rural and urban areas at each PHUs which may have increased the risk of Mpox throughout Ontario. Although, higher proportion ethnic concentration or visible minority groups may be significantly located in urban areas and higher material deprivation greater in rural areas across the PHUs in Ontario (33). We found higher Mpox incidence rates in PHUs with higher racial population (visible minority or ethnic concentration) which is the most important predictor for high-risk Mpox incidence across PHUs in Ontario. This is in line with other recent Mpox studies (56), indicating that PHUs with dense populations, and high migration rate in Ontario, are at high risk.

This result indicates that human population density is an important variable in Mpox distribution, which may be caused by increased human-environment interaction among large urban centers. Areas with the greater ethnic concentration had the highest risk of Mpox. Previous studies looking at the risk of other diseases like COVID-19 among immigrants and the socially deprived population in Ontario found elevated cases of the disease in some immigrant and socially deprived groups and lower rates among others (57–62). This could be attributed to immigrants or visible minority groups living in crowded households, lower income areas and working in an environment where there is frequent physical contact. The results indicate that disease spread in Ontario is associated with health inequity, meaning that some individuals may be infected by diseases because they cannot reach their full health potential and are disadvantaged from attaining it because of their race, gender, age, socioeconomic status or other socially determined factors. However, international connections of migrants and racial minorities can be significant in impacting both individual lives and broader societal dynamics, often participating actively in the economy, starting businesses, filling essential job roles, and contributing to economic growth through their labor and entrepreneurial activities.

The result of this study collaborates previous studies on Mpox, suggesting that younger and middle-aged adults were more vulnerable to Mpox infection, especially with male gender (22, 60). Therefore, controlling population activities and monitoring public health management and vaccination in areas with high population density can effectively control the spread of Mpox virus must target younger age groups especially the male gender. At the same time, it is necessary to improve the public’s awareness of the Mpox virus, understanding the changing epidemiology of the disease is also the key to formulating prevention and control strategies. Avoiding human interaction in areas where suspicious host activities are frequent will reduce the likelihood of introducing Mpox into the scope of human activities and if Mpox is not introduced repeatedly, the infection will eventually stop occurring in Toronto and other high-risk PHUs.

Surprisingly, we found that after adjusting for other variables in the GPR model, the material deprivation was found to be a negative predictor to Mpox incidence in Ontario but not significant, suggesting that higher Mpox incidence occurred in PHUs with better socioeconomic indicators, however, household instability was found to be a significant risk factor for Mpox. This implies that Mpox incidence occurred in a highly socioeconomic PHUs hence related to population density and personal interaction. Despite the progress made towards reducing health inequality in Ontario, there are certain Ontario PHUs that may not be benefiting fully from Ontario’s health system improvements. It could be explained that those with low educational levels and low-income individuals typically move into areas where there is a predominant dependence on welfare assistance and other health-related services. Motives for moving regularly by such persons may be attributed to the unavailability of income and health assistance services in addition to poor prospects for employment for such a category of persons and cheaper housing rates. Similarly, findings from other studies indicated that individual income levels alone did not affect Mpox disease infection, but the relative location of residence did (62, 63).

### Study’s strength and Limitation

There are several limitations to our study. First, we used a limited number of variables based on publicly available data at the PHU level. While Ontario marginalized index used various census variables for 2021 to define dimensions of marginalization, some details are not well captured like the issue of racism, and health literacy. Similarly, given that ON-Marg index factors only explain a portion of Mpox incidence, it is important to consider other environmental, behavioral, and biological factors that impact a population’s risk of contracting Mpox. Furthermore, this study was not structured to establish causal relationships, nor does it consider all the factors that might contribute to a causal pathway. However, our study is first to use a comprehensive marginalization index as social determinants of health to better understand the incidence of Mpox in Ontario.

### Conclusion

This study has demonstrated geographical distribution and the impact of marginalization on Mpox incidence across the PHUs in Ontario. Our study showed that most Mpox incidences in Ontario occurred in PHUs with better socioeconomic indicators and higher population density (higher proportion racial and ethnic minority groups). Additionally, household dwelling (residential instability) was found to be associated with increased risk of Mpox. Measures to enhance Mpox diagnosis and promote health equity among socioeconomically vulnerable populations, including individuals from racial and ethnic minority backgrounds, need to be put into action. This study offers valuable insights for resource allocation and policymaking for Mpox control and prevention. Further studies and policy intervention will be invaluable not only to identify and support socially marginalized populations in Ontario but also to address possible racial issues on a large scale in context of Mpox control and resource allocation.

## Data Availability

The data underlying the results presented in the study are available from the Integrated Public Health Information System (iPHIS) of Public Health Ontario (PHO): https://www.publichealthontarioca/en/Diseases-and-Conditions/Infectious-Diseases/Vector-Borne-Zoonotic-Diseases/mpox, and https://www.12statcangcca/census-recensement/2021/geo/sip-pis/boundary-limites/index2021-engcfm?year=21

https://www.publichealthontario.ca/en/Data-and-Analysis/Health-Equity/Ontario-Marginalization-Index

https://www.12statcangcca/census-recensement/2021/dp-pd/prof/indexcfm?Lang=E

https://www.12statcangcca/census-recensement/2021/geo/sip-pis/boundary-limites/index2021-engcfm?year=21

https://www.publichealthontarioca/en/Diseases-and-Conditions/Infectious-Diseases/Vector-Borne-Zoonotic-Diseases/mpox

https://health-infobasecanadaca/mpox/%23detailedCases

## Acknowledgments

W.A.W acknowledges financial support from the NSERC Discovery Grant (Appl No.: RGPIN-2023-05100).

